# Cardiac contractility index identifies systolic dysfunction in preserved ejection fraction heart failure

**DOI:** 10.1101/2022.11.22.22282605

**Authors:** Sam Straw, Charlotte Cole, Oliver Brown, Judith Lowry, Maria F Paton, Ruth Burgess, Michael Drozd, Thomas A Slater, Samuel D Relton, Richard M Cubbon, Eylem Levelt, Klaus K Witte, Mark T Kearney, John Gierula

**Author notes:** **Corresponding author:** Dr John Gierula. Leeds Institute of Cardiovascular and Metabolic Medicine, LIGHT Building, University of Leeds, Clarendon Way, Leeds, LS2 9JT, UK. denotes joint first authorship (Sam Straw, Charlotte Cole).

## Abstract

**Background:** Left ventricular ejection fraction (LVEF) has well-known limitations including modest reproducibility, load dependence, and representation of the percentage change in left ventricular (LV) volume rather than myocardial contractility. We aimed to assess the prognostic value of systolic blood pressure: indexed left ventricular end-systolic volume ratio, or ‘cardiac contractility index’ (CCI).

**Methods:** We conducted a prospective cohort study in 728 unselected individuals newly diagnosed with chronic heart failure. We divided patients into tertiles of LVEF and CCI, and also divided those with heart failure with reduced (HFrEF) or preserved ejection fraction (HFpEF) by the median value of CCI (4.43mmHg/ml/m^2^) into four groups. Mortality rates for CCI and LVEF as continuous variables were assessed using unadjusted and adjusted Poisson regression models.

**Results:** There was a modest, positive correlation between LVEF and CCI (r=0.70 [0.66-0.74], R^2^ 0.49; *p*<0.0001), although the latter was distributed widely for any given value of LVEF, especially for those with HFpEF. We observed distinct clinical characteristics across tertiles of both LVEF and CCI, with an inverse relationship with conventional markers of risk including N-terminal B-type natriuretic peptide (*p*<0.001 in both comparisons). There was a clear relationship between tertiles of CCI and all-cause mortality risk, which was less evident when patients were divided by LVEF. When modelled as continuous variables there was a curvi-linear relationship between all-cause mortality rates and CCI, but the relationship between LVEF and mortality risk was more complex, with no clear association across a wide range from 25-55%. In models including relevant covariates, the association between LVEF and mortality was no longer evident except for those with LVEF 60% (relative to 50%) but remained evident for all specified values of CCI. Patients with HFpEF and CCI below the median value had an all-cause mortality risk ∼40% higher than those with CCI above median (*p*<0.001), similar to those with HFrEF.

**Conclusions:** CCI is a non-invasive, relatively afterload independent measure left ventricular contractility which provided additional prognostic information beyond conventional assessment by LVEF. Furthermore, CCI was able to reclassify around a third of patients with HFpEF, and these patients had distinct characteristics and a worse prognosis.

**Clinical perspective:** 

**What’s new?:**

- In an unselected population with chronic heart failure, cardiac contractility index (CCI) provided better prognostic accuracy than left ventricular ejection fraction.
- CCI was able to reclassify around a third of patients with a preserved ejection fraction who had evidence of reduced left ventricular contractility, and these patients had distinct characteristics and all-cause mortality risk similar to those with a reduced ejection fraction.

**What are the clinical implications?:**

- CCI is a simple, relatively afterload independent measure of left ventricular contractility, which utilises data already part of a standard echocardiographic assessment.
- The identification of subtle or concomitant systolic dysfunction in heart failure with a preserved ejection fraction may help better define risk and refine the phenotypic classification of this heterogenous group.

## Introduction

### Background

Chronic heart failure is a clinical syndrome characterised by breathlessness, fatigue, frequent hospitalisation, and premature death.^1^ Current recommendations classify patients by left ventricular ejection fraction (LVEF), the most commonly reported measure of systolic function. Around half of people with heart failure have a preserved ejection fraction (HFpEF), and these individuals have similar symptoms, impairments in quality of life, hospitalisation rates, and mortality risk as those with heart failure with a reduced ejection fraction (HFrEF).^2^ As a measure of systolic dysfunction, LVEF has well-known limitations including modest reproducibility, load dependence, and representation of the percentage change in left ventricular (LV) volume rather than myocardial contractility.^3^

Although diastolic dysfunction has been proposed as a key mechanism underpinning the pathophysiology of HFpEF, the presence of subtle or concomitant systolic dysfunction as assessed by strain imaging in those with LVEF ≥50% has been previously suggested.^4^ Simply classifying patients as having an ejection fraction which is ‘normal’ may offer limited insight and risk misclassifying those who have any degree of systolic dysfunction. The ability to more easily identify individuals with HFpEF who actually have concomitant systolic dysfunction could help further refine its phenotypic classification, stratify risk, and identify potential to derive benefit from disease modifying pharmacological therapies.

LV end-systolic pressure to LV end-systolic volume index ratio, or ‘cardiac contractility index’ (CCI), is a non-invasive measure of LV contractility, validated against invasive haemodynamic studies.^5,6^ By incorporating LV end-systolic pressure, CCI is relatively independent of afterload and may therefore more accurately reflect LV contractile force. CCI has been used as a surrogate endpoint in randomised controlled trials assessing pharmacological and device-based therapies in HFrEF, although its prognostic significance in heart failure is unknown.^7,8^

### Aims

In this study we firstly sought to evaluate whether CCI was associated with all-cause mortality risk in an unselected population with chronic heart failure. Secondly, we aimed to determine its prognostic accuracy compared to LVEF. And, thirdly, we assessed whether CCI could reclassify patients with HFpEF and if these patients had a distinct phenotype and mortality risk.

## Methods

### Patient population

The prospective evaluation of the diagnostic efficacy of the 2010 United Kingdom National Institute for Clinical Excellence guidelines on Chronic Heart Failure (NICE-CHF) is an observational cohort study including people newly referred to secondary care for suspected heart failure.^9^ Consecutive unselected patients from a primary care catchment of over 750,000 people between May 2012 and May 2013, who had signs and/or symptoms of chronic heart failure as well as elevated natriuretic peptides (N-terminal pro B-type natriuretic peptide [NT-proBNP] ≥125pg/L), were included.

### Study procedures

Patients were evaluated in a secondary care specialist heart failure clinic. Upon arrival, demographic details, medical history, and currently prescribed medications were recorded. Height and weight were measured, and a venous blood sample was taken and tested for full blood count, electrolytes, and assessment of renal and liver function. NT-proBNP had previously been measured at the point of referral using samples collected in primary care, which were analysed at our institution using the Immulite 2000 assay (Siemens Healthcare Diagnostics, Camberley, UK) which has an inter-batch coefficient variation of 8.9% at 350pg/mL and 5.9% at 4100pg/ml. A standard 12-lead electrocardiogram was recorded at 25mm/s and two-dimensional transthoracic echocardiography was performed.

### Echocardiography analyses

To determine CCI, systolic blood pressure was used as a surrogate of LV end-systolic pressure as previously described.^10^ This was measured using a standard sphygmomanometer placed on the patient’s right arm whilst in a supine position immediately prior to echocardiography. Echocardiographic images were obtained by senior cardiac sonographers according to recommendations of the American Society of Echocardiography and European Association of Cardiovascular Imaging.^11^ Images were then sent to digital storage media and, for the present analysis were analysed offline using Medcon (McKesson Cardiology, Irving TX, USA) by two senior accredited cardiac sonographers who were blinded to patient characteristics and measurements of NT-proBNP. Where endocardial border definition allowed, LV end-diastolic and end-systolic volumes were measured in apical two and four-chamber views using the biplane method of disks, and indexed for body surface area using the Mosteller equation.^12^ CCI was calculated by dividing systolic blood pressure by LV end-systolic volume indexed to body surface area.

### Patient classification and ascertainment of outcomes

All patients in the present study had signs and/or symptoms of chronic heart failure and elevated natriuretic peptides (N-terminal B-type natriuretic peptide ≥125pg/mL). We categorised patients according to the most recent European Society of Cardiology guidelines.^1^ For simplicity we categorised all patients with LVEF <50% as having HFrEF, and those with LVEF ≥50% as well as relevant structural heart disease (either left atrial dilatation, LV hypertrophy) or diastolic dysfunction, as having HFpEF.^1^ Patients without these echocardiographic features, including those in whom symptoms were attributable to significant valvular disease were excluded. We divided patients into tertiles based on measures of LV function, to determine the clinical characteristics and outcomes of these groups. The ranges for tertiles one, two and three of LVEF were <46.2%, 46.2-55.1% and >55.1%; the ranges for tertiles one, two and three of CCI were <3.65mmHg/ml/m^2^, 3.65-5.34mmHg/ml/m^2^ and >5.34mmHg/ml/m^2^. We then subdivided patients into four groups according to whether they were classified as having HFrEF or HFpEF and whether CCI was above or below the median value of 4.43mmHg/ml/m^2^. The primary outcome was all-cause mortality according to LVEF and CCI. Vital status data were collected using linked Hospital Episode Statistics and Office of National Statistical mortality data which were available for all patients with final censorship in April 2019.

### Statistical analysis

This was an observational study of consecutive cases of newly diagnosed heart failure during one year, and the sample size was not prespecified. Normality of distribution was confirmed using skewness tests. Continuous variables are presented as mean ± standard deviation if normally distributed, or as median (interquartile range) if non-normally distributed, with discrete variables presented as number (percentage). Groups were compared using t-tests or one-way analysis of covariance for normally distributed continuous data, Mann-Whitey or Kruskal-Wallis H tests for non-normally distributed data, and Pearson χ^2^ test for categorical variables. Interobserver variability for LV end-diastolic and end-systolic volumes were compared in a random sample of 10% of patients assessed by both observers by inter-observer correlation coefficient and displayed by Bland-Altman plots. We determined the association between LVEF and CCI by Pearson’s correlation. We plotted Kaplan Meier curves to illustrate all-cause mortality rates, with significance testing between groups determined by log-rank test.

We found the proportional hazards assumptions were not valid for LVEF and CCI, we therefore estimated mortality rates ratios (IRR) using Poisson regression models. Exposure time was modelled, and we chose four knots for both variables as these provided the best fit assessed by the Akaike and Bayesian information criterion scores. Models including cubic splines with three, four or five knots and first and degree fractional polynomials were compared and found to provide less robust fit. IRRs estimated for LVEF and CCI pertain to specific points (LVEF 20, 30, 40, and 60% compared with 50%, and CCI 2, 4, 6 and 8mmHg/ml/m^2^ compared with 4.43mmHg/ml/m^2^ which was the median value). Covariates included in a multivariable Poisson regression model were age, sex, ischaemic heart disease, diabetes mellitus, hypertension, systolic blood pressure, heart rate, haemoglobin, creatinine, albumin and NT-proBNP, in which non-normally distributed continuous data were log10 transformed. All tests were two-sided and statistical significance was regarded as *p*<0.05. Statistical analyses were done using Stata/MP (version 16.1, StataCorp LLC, College Station, TX, USA) and PRISM (version 9, GraphPad Software Inc, San Diego, CA).

### Ethical considerations

The United Kingdom Health Research Authority provided ethical approval for the study through a Section 251 application reviewed by the Confidential Advisory Group. Approval through a Section 251 application allows individual patient data to be used for health service improvement and waives the requirement for individual patient consent (CAG8-03(PR1)/2013). Appropriate data safeguards were in place and the study complied with the principles outlined in the Declaration of Helsinki.

## Results

### Baseline characteristics of the study population

Between May 2012 and May 2013, a total of 982 patients who had suspected heart failure and NTpro-BNP ≥125pg/L were referred for evaluation in secondary care. Of these, 182 did not fulfil diagnostic criteria for chronic heart failure (Supplementary Table 1), whilst for a further 72 patients, calculation of CCI was not possible due to either insufficient endocardial definition or missing height, weight, or systolic blood pressure. The final dataset therefore consisted of 728 patients who had a mean age of 82.6 ± 9.2 years, of whom 398 (54.7%) were male. The inter-observer correlation coefficients for LV end-diastolic and end-systolic volumes were 0.99 (95% CI 0.99-1.00) and 0.99 (95% CI 0.99-1.00), respectively (Supplementary Figure 1). Across the entire dataset the mean LVEF was 48.2 ± 11.6% (range 12.7-68.9%) with 293 (40.2%) classified as having HFrEF and 435 (59.8%) as having HFpEF. The mean CCI was 4.55±1.9 mmHg/ml/m^2^ (range 0.71-11.3mmHg/ml/m^2^) (Supplementary Figure 2).

### Clinical characteristics according to LVEF and cardiac contractility index

We divided patients into tertiles of LVEF and CCI to determine differences in the demographic and clinical characteristics between these groups (Table 1). Patients within the lowest tertiles of both LVEF and CCI were more often male, more frequently had ischaemic heart disease and diabetes mellitus, and were less likely to have hypertension. They also had on average greater conventional markers of disease severity including lower systolic blood pressure, higher heart rate, and worse renal function.

There was an inverse relationship between NTpro-BNP and both LVEF and CCI. Median NTpro-BNP was 655 (345-1344) pg/mL, 1003 (503-2113) pg/mL and 2119 (810-4827) pg/mL for tertiles one, two and three of LVEF, respectively; and 715 (348-1362) pg/mL, 921 (486-1759) pg/mL and 2310 (933-5155) pg/mL for tertiles one, two and three of CCI, respectively (*p*<0.0001 for trend in both comparisons) (Figure 1). Despite this, the proportion of patients with New York Heart Association class III/IV symptoms was similar across tertiles of LVEF and CCI. Aside from the variables from which these groups were derived (systolic blood pressure and indexed LV volumes), the pattern of these associations was similar across tertiles whether patients were divided by LVEF or CCI.

**Figure 1.**
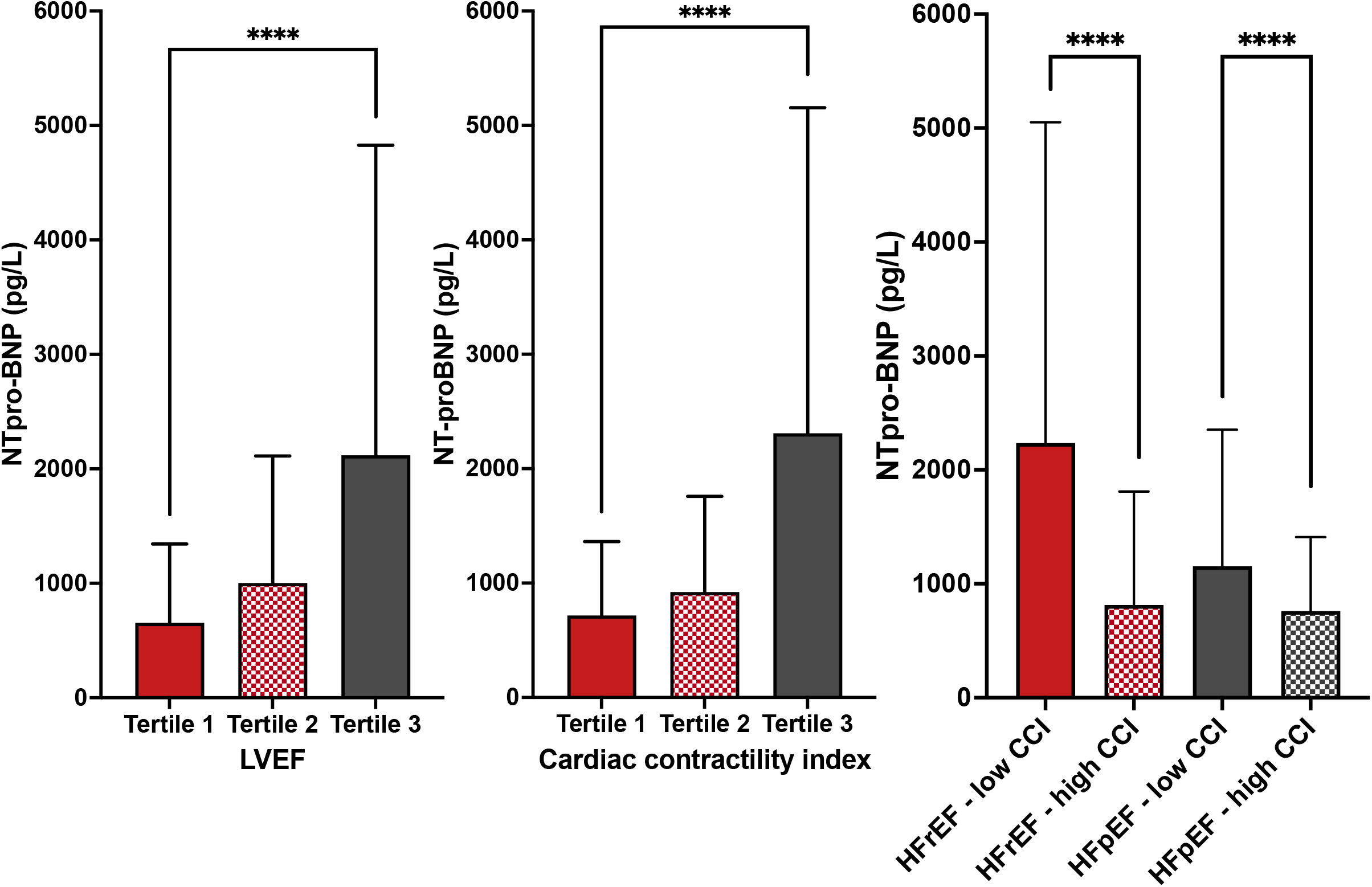
Bar charts showing levels of NT-proBNP between groups.

### Relationship between LVEF and cardiac contractility index

Although there was a modest, positive correlation between LVEF and CCI (r=0.70 [0.66-0.74], R^2^ 0.49; *p*<0.0001), the latter was distributed widely for any given value of LVEF, especially evident for those with a preserved ejection fraction (Figure 2). To explore the relationship between LVEF and CCI further, we divided patients with HFrEF and HFpEF according to median CCI (≤ or >4.43mmHg/ml/m^2^) into four groups. In the HFrEF group, 232 (79.2%) had low contractility and 61 (20.8%) high contractility; of patients with HFpEF, 132 (30.3%) had low contractility and 303 (69.7%) had high contractility.

**Figure 2.**
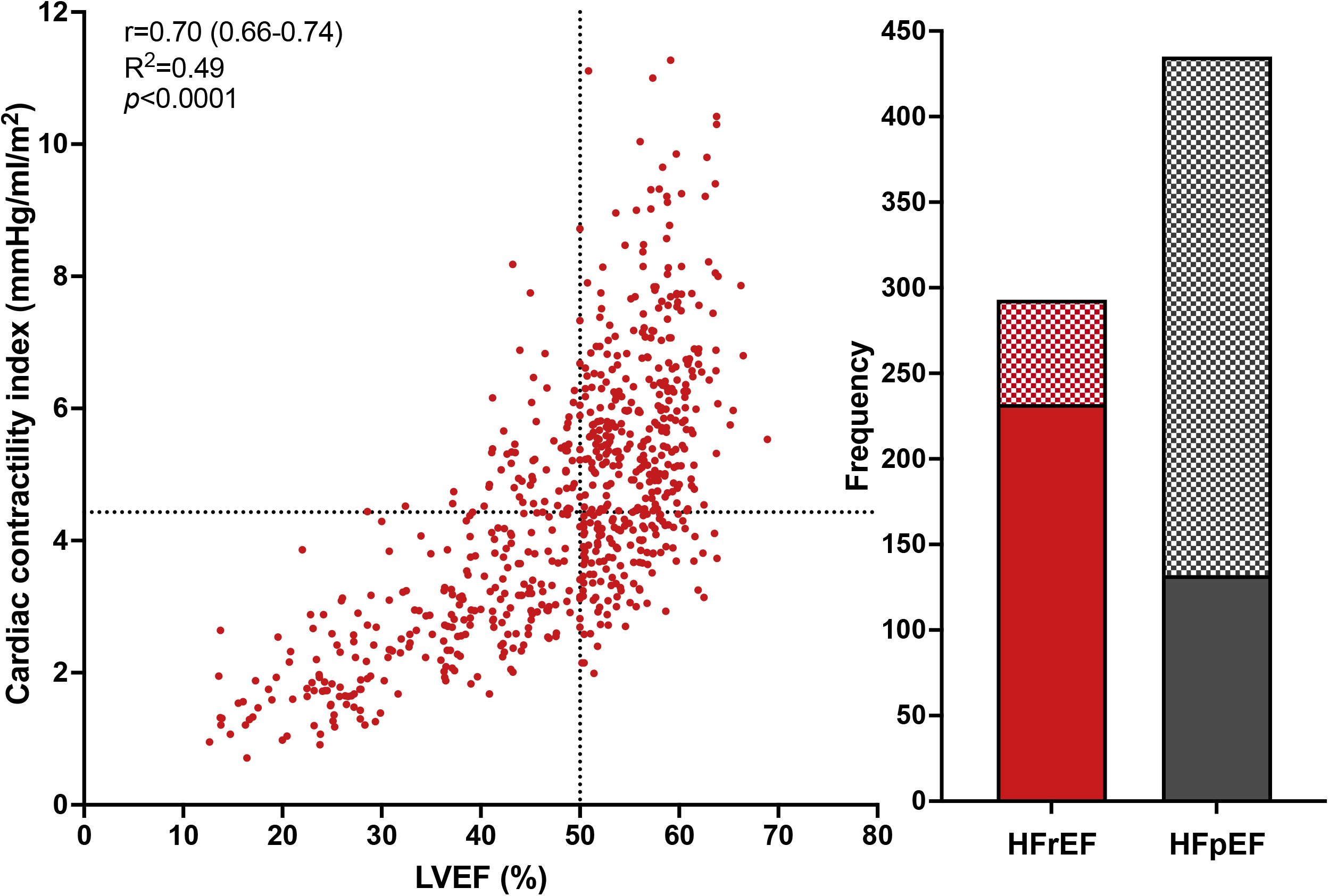
Scatter plot of LVEF and CCI and bar charts showing the proportion of patients with HFrEF and HFpEF who had low or high CCI.

The clinical characteristics of patients with HFrEF and HFpEF divided according to median CCI are displayed in Table 2. Patients classified as having HFpEF were on average older, less likely to be male, have ischaemic heart disease or diabetes mellitus and more likely to have hypertension and were receiving lower doses of loop diuretics compared to patients with HFrEF. Within patients classified as having HFpEF, those with low cardiac contractility index were more often male, had ischaemic heart disease and had other markers of risk including lower serum haemoglobin, worse renal function and lower serum albumin compared to those with HFpEF and high contractility. Across these groups, we observed that median NT-proBNP was higher in those with low CCI, regardless of whether they were classified as having HFrEF (2235 [788-5052] and 813 [450-1810] pg/mL) or HFpEF (1153 [503-2353] pg/mL and 761 [401-1409] pg/mL) (Figure 2).

### Associations with outcomes

During a median follow-up of 5.9 (2.9-9.0) years, a total of 491 (67.4%) patients died. We observed incrementally lower mortality risk across tertiles of CCI, whereas for LVEF, mortality risk was similar comparing patients in tertiles one and two (Figure 3). All-cause mortality risk was more clearly distinguished between CCI groups than the groups as defined by LVEF regardless of whether divided into two groups, tertiles or quartiles (Supplementary Figure 3).

**Figure 3.**
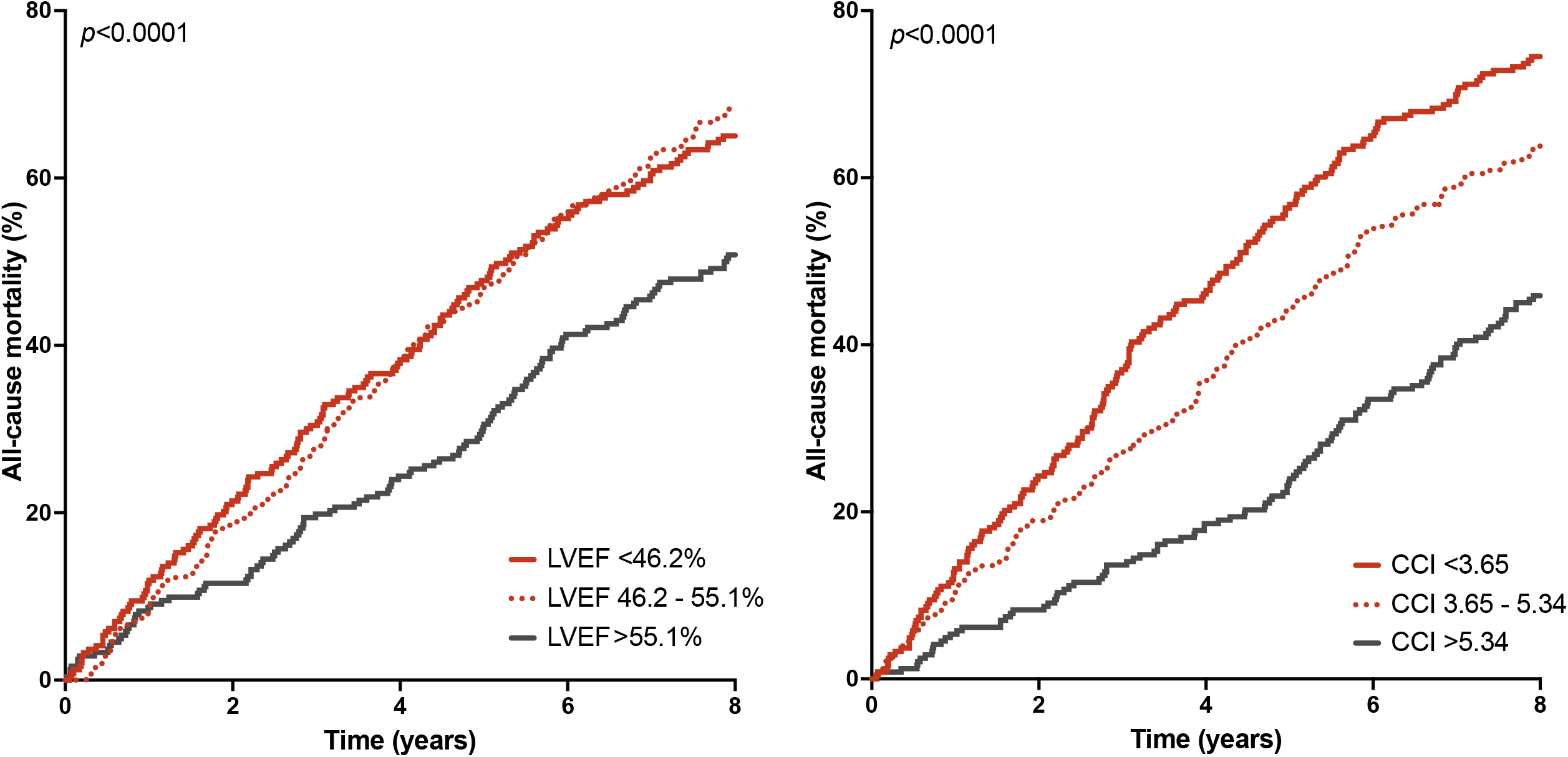
Kaplan-Meier plot of all-cause mortality divided by tertiles of LVEF and CCI.

We further evaluated the relationship between mortality and LVEF or CCI by modelling them as continuous variables using restricted cubic splines. We observed a curvi-linear relationship with all-cause mortality risk for CCI, with significantly higher or lower mortality rates across a broad range below or above the median, respectively (Figure 4). The relationship with LVEF was more complex, with no clear association with mortality rates across a wide range from 25 to 55% (Figure 3). Furthermore, in a model including relevant covariates (Table 3), the association between LVEF and mortality was no longer evident except for those with the highest LVEF in whom the rate was lower (LVEF 60% IRR 0.69 [0.54-0.88], relative to LVEF 50%). In contrast, the association with all-cause mortality rate remained evident for all specified values of CCI, even after adjusting for prognostically important covariates.

**Figure 4.**
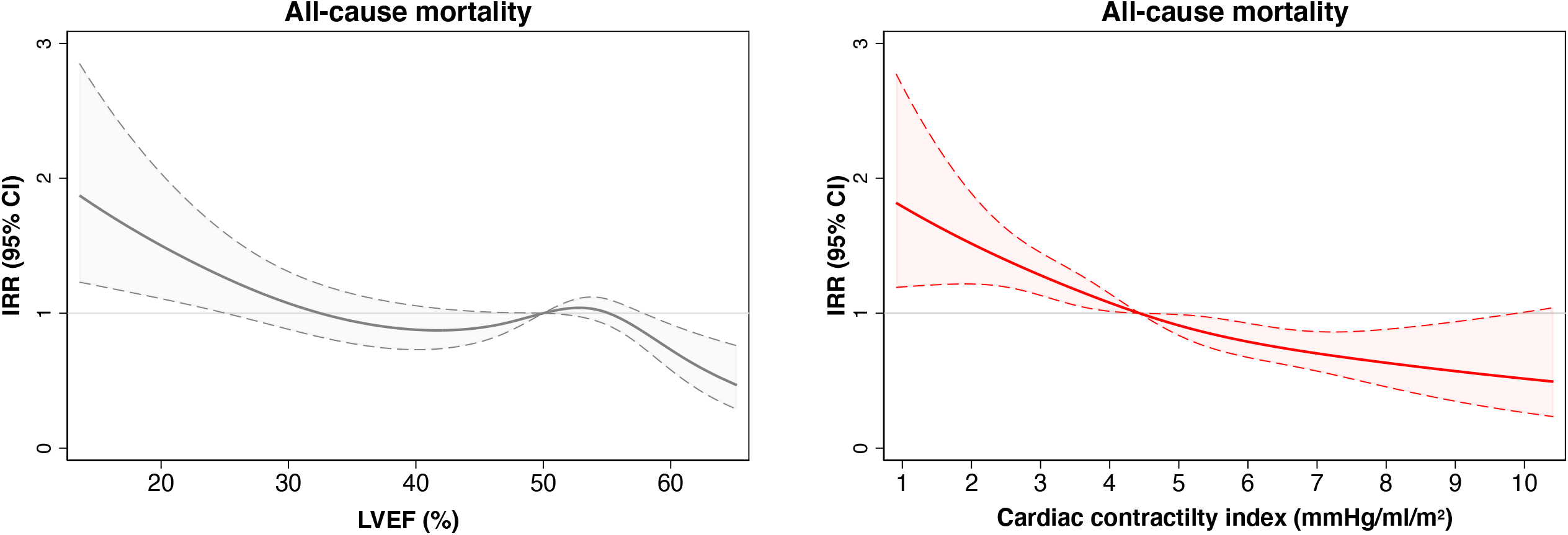
Restricted cubic splines displaying incidence rate ratios and 95% confidence intervals of all-cause mortality across LVEF and CCI.

When these two measures of LV systolic function were combined, we observed the all-cause mortality risk was similar for patients classified as having HFrEF, regardless of whether CCI was above or below the median value (*p*=0.096). However, patients with HFpEF and below median CCI, had an all-cause mortality risk ∼40% higher than those patients with CCI above median (*p*<0.001), which was similar to people with HFrEF (Figure 5).

**Figure 5.**
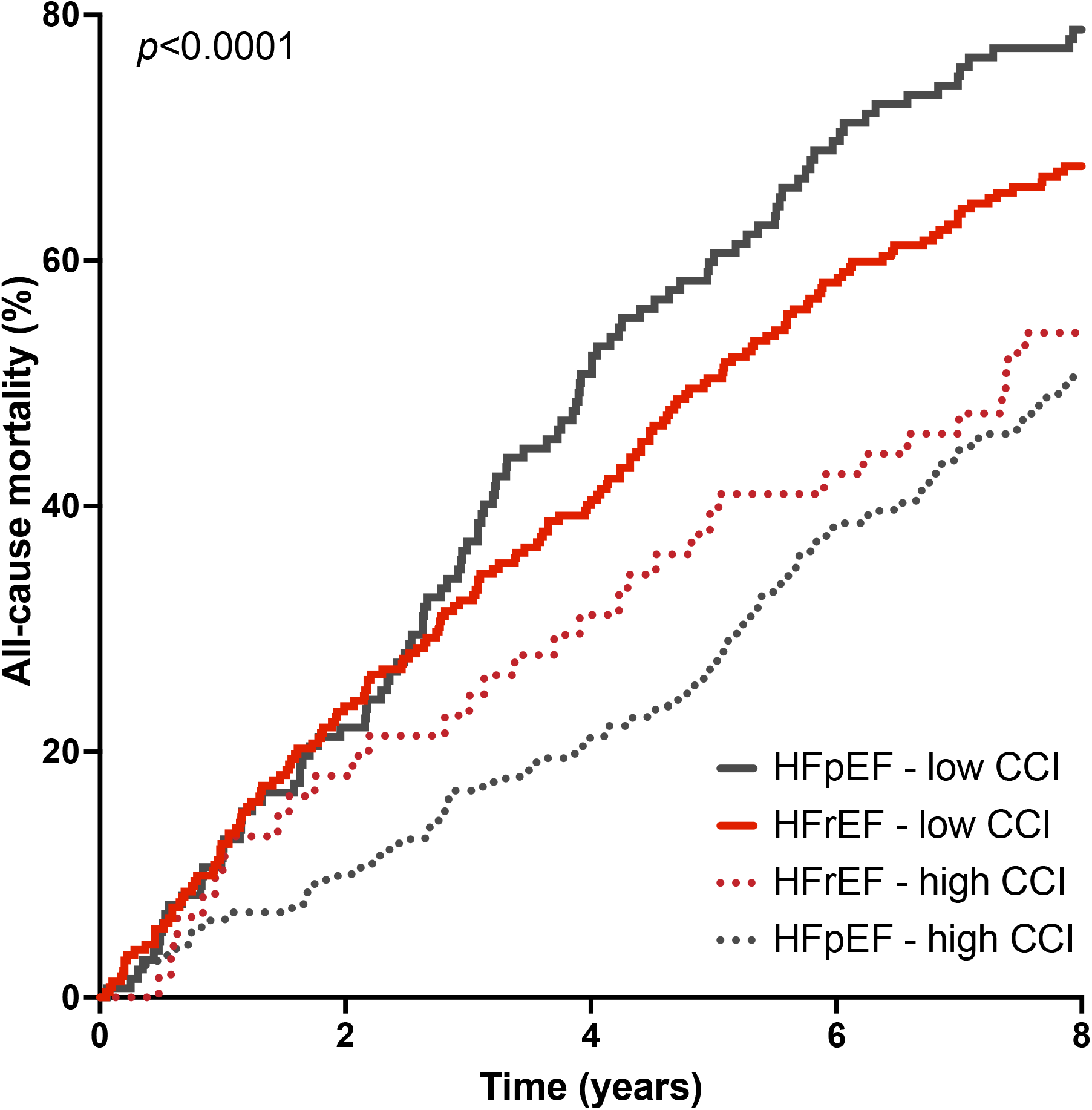
Kaplan-Meier plot of all-cause mortality in patients with HFrEF and HFpEF who had CCI below or above the median value.

## Discussion

### Principal findings

Our analysis of consecutively presenting patients with *de novo* chronic heart failure has four novel findings. First, we observed a broad range of LV contractility, especially amongst patients classified as having a preserved ejection fraction. Second, we observed a clear relationship between CCI and mortality, which remained evident in a model including conventional markers of risk. Third, the association with all-cause mortality was less clear when patients were classified according to LVEF and was not evident across a broad range of values in adjusted analyses. Fourth, when patients with HFpEF were reclassified as having low CCI, the all-cause mortality risk was similar to those with HFrEF, despite distinct clinical characteristics. Taken together, these data suggest CCI may help further refine the phenotypic classification of HFpEF by identifying patients with subtle systolic dysfunction who have a worse prognosis.

### Left ventricular ejection fraction as an imperfect but essential tool in chronic heart failure

First described over six decades ago,^13^ LVEF is a simple measure of LV systolic function which can be applied across different imaging modalities. Much of current practice is anchored to this simple variable, primarily as landmark trials supporting current guideline-directed therapies enrolled patients below arbitrary thresholds of LVEF who were perceived to be at the highest risk.^14,15^ For patients with HFrEF, four classes of medications targeting the neurohormonal maladaptations of the syndrome are proven to reduce hospitalisations and improve survival.^16^ However, with the possible exception of sodium-glucose co-transporter 2 inhibitors (for which there is conflicting evidence),^17,18^ the benefits of these agents are attenuated for those with higher LVEF.^19,20^ Moreover, for the overall population with HFpEF no therapies have been shown to improve survival, with the positive results of recent trials largely driven by reductions in heart failure hospitalisations.^21,22^ As a result, in clinical practice LVEF persists as an imperfect but necessary tool for diagnosis, risk stratification, and to determine in whom currently available therapies should be applied.

### Limitations of left ventricular ejection fraction

Whilst LVEF remains central to our understanding of heart failure, its relative simplicity comes with several well-known limitations, including poor intra and inter-observer reproducibility, depending on the methods and experience of observers.^23^ Furthermore, whilst in the acute setting (for example following myocardial infarction) reduced LVEF may be truly reflective of reduced LV contractility due to the loss of cardiomyocytes, over time LV dilatation means LVEF in chronic heart failure is principally reflective of remodelling in response to loading conditions, and therefore poorly reflects myocardial contractile force.^24^ This might be acceptable in conditions such as dilated cardiomyopathy, in which increases in end-diastolic volume parallels reductions in systolic function. In this setting, where stroke volume is initially preserved, dilatation is reflected by a declining LVEF which therefore provides a good approximation of systolic function.^24^ However, in other disease states such as restrictive or infiltrative cardiomyopathies, even when myocardial shortening is impaired, there is no resultant increase in LV volumes, such that the measured LVEF is maintained even though systolic function is compromised. In our patients, concomitant systolic dysfunction, determined by low CCI in those with HFpEF was associated with a worse prognosis, independent of clinical characteristics, which were distinct from those with HFrEF.

### The potential advantages of cardiac contractility index

A future without LVEF to guide therapy seems improbable, but additional indices of systolic performance which could be incorporated into routine clinical practice may help guide care. The current research landscape of alternative imaging modalities is dominated by speckle tracking techniques such as myocardial strain and strain rate. Myocardial strain has been shown to better reflect systolic dysfunction amongst patients with HFpEF,^25^ and global longitudinal strain is independently associated with mortality risk in hospitalised patients, providing prognostic information not revealed by LVEF.^3^ However, barriers exist to its more widespread adoption such as variation between vendors and as yet, no agreed definition of normal ranges. Additionally, there are limited data on the effects of loading conditions on these measurements, whilst the most commonly reported metric, global longitudinal strain, assesses only longitudinal function meaning circumferential and radial dysfunction may be overlooked.

On the other hand, CCI is a simple parameter, with the advantage of being relatively load independent, thereby reflecting changes in LV contractility.^27^ As originally described, CCI requires a measurement of LV volume and pressure at end-systole by invasive ventriculography. However the use of non-invasive systolic blood pressure obtained by standard sphygmomanometer as a surrogate for end-systolic pressure and has been validated against invasive measurements,^10^ is already part of a standard echocardiographic assessment, and, in our patients outperformed a conventional assessment of systolic function by LVEF. Low systolic blood pressure is commonly observed in those with advanced systolic heart failure, and these patients seem to derive the greatest benefits from therapies targeting LV contractility.^26^

When applied to the current dataset, the relationship with all-cause mortality risk and CCI was curvi-linear and more robust than for LVEF, for which the association with mortality was not evident throughout most of its range. Moreover, by applying CCI to those with HFpEF we were able to show that around a third of patients had reduced contractility, and that these patients had an all-cause mortality risk similar to those with HFrEF. Although dividing patients by the median value of CCI was arbitrary, with as yet no data describing normal values in healthy populations, by doing so we were able to identify a subgroup of people classified as having a normal ejection fraction who were phenotypically distinct and had worse outcomes.

### Strengths and limitations

Our analysis includes consecutively referred patients from a prospective study, representative of real-world populations of patients with CHF, encompassing a broad spectrum of LVEF with long-term mortality data.^28^ Some limitations should be noted. Firstly, this was an observational study conducted in a single centre which may limit generalisability, although the diverse characteristics of the area served by our centre and the inclusion of consecutive patients referred to our service mitigates against this.^29^ Secondly, the lack of longitudinal imaging data means we cannot determine whether patients with HFpEF went on to develop overt systolic dysfunction (LVEF <50%) in the future and whether low CCI predicted this. Third, normal values of CCI have not been defined and is it therefore unknown how thresholds defined by median values in the present dataset align with those from healthy populations. Fourth, we did not assess the prognostic value of other measurements of LV systolic function such as myocardial strain which may further refine phenotypic classification of patients with heart failure, although the simplicity of CCI means it could be applied retrospectively to datasets in which myocardial strain or strain-rate were assessed.

### Conclusion

Our findings suggest CCI may provide additional prognostic information, especially for those patients with heart failure and an ejection fraction currently classed as normal. For these patients, the identification of unappreciated systolic dysfunction may help better define risk or refine the phenotypic classification of this heterogenous group. These data could also help refine the inclusion criteria of future randomised controlled trials, potentially allowing us to establish effective therapies for HFpEF patients stratified by their LV contractility. In the meantime, its simplicity means this variable could be easily applied to existing datasets in order to identify who may have derived benefits from therapies in which the overall population did not.

## Data Availability

The datasets generated and/or analysed during the current study are not publicly available due to the inclusion of potentially identifiable information but are available from the corresponding author upon reasonable request.

## Acknowledgements

SS, MD, JEL, RMC and MTK are supported by the British Heart Foundation. JG and MFP are supported by the National Institute of Health Research. EL is supported by the Wellcome Trust. The authors acknowledge the support of the National Institute of Health Research Leeds Cardiovascular Clinical Research Facility.

## Authorship

CC, JEL, MFP, TAS, RMC, MTK, KKW and JG collected the data. SS, OB and RMC analysed the data. SS produced the first draft of the manuscript. All other authors provided critical revision.

## supplementary figure legends

<u>supplementary Figure 1</u>

Bland-Altman plots of inter-observer variability of LV end-diastolic and end-systolic olumes.

<u>supplementary Figure 2</u>

Frequency distribution plots of LVEF and cardiac contractility index.

<u>supplementary Figure 3</u>

Kaplan-Meier plots of all-cause mortality divided by median values or into quartiles of VEF and CCI.

## References

1. McDonagh TA, Metra M, Adamo M, et al. 2021 ESC Guidelines for the diagnosis and treatment of acute and chronic heart failure. Eur Heart J. 2021;42(36):3599–3726.

2. Shah KS, Xu H, Matsouaka RA, et al. Heart Failure With Preserved, Borderline, and Reduced Ejection Fraction: 5-Year Outcomes. J Am Coll Cardiol. 2017;70(20):2476–2486.

3. Park JJ, Park JB, Park JH, Cho GY. Global Longitudinal Strain to Predict Mortality in Patients With Acute Heart Failure. J Am Coll Cardiol. 2018;71(18):1947–1957.

4. Kraigher-Krainer E, Shah AM, Gupta DK, et al. Impaired systolic function by strain imaging in heart failure with preserved ejection fraction. J Am Coll Cardiol. 2014;63(5):447–456.

5. Ginzton LE, Laks MM, Brizendine M, Conant R, Mena I. Noninvasive measurement of the rest and exercise peak systolic pressure/end-systolic volume ratio: a sensitive two-dimensional echocardiographic indicator of left ventricular function. J Am Coll Cardiol. 1984;4(3):509–516.

6. Bombardini T, Correia MJ, Cicerone C, Agricola E, Ripoli A, Picano E. Force-frequency relationship in the echocardiography laboratory: a noninvasive assessment of Bowditch treppe? J Am Soc Echocardiogr. 2003;16(6):646–655.

7. Martens P, Dupont M, Dauw J, et al. The effect of intravenous ferric carboxymaltose on cardiac reverse remodelling following cardiac resynchronization therapy-the IRON-CRT trial. Eur Heart J. 2021;42(48):4905–4914.

8. Gierula J, Lowry JE, Paton MF, et al. Personalized Rate-Response Programming Improves Exercise Tolerance After 6 Months in People With Cardiac Implantable Electronic Devices and Heart Failure: A Phase II Study. Circulation. 2020;141(21):1693–1703.

9. Straw S, Cole CA, McGinlay M, et al. Guideline-directed medical therapy is similarly effective in heart failure with mildly reduced ejection fraction. Clin Res Cardiol. 2022.

10. Haedersdal C, Madsen JK, Saunamaki K. The left ventricular end-systolic pressure and pressure-volume index. Comparison between invasive and auscultatory arm pressure measurements. Angiology. 1993;44(12):959–964.

11. Lang RM, Badano LP, Mor-Avi V, et al. Recommendations for cardiac chamber quantification by echocardiography in adults: an update from the American Society of Echocardiography and the European Association of Cardiovascular Imaging. J Am Soc Echocardiogr. 2015;28(1):1–39 e14.

12. Mosteller RD. Simplified calculation of body-surface area. N Engl J Med. 1987;317(17):1098.

13. Folse R, Braunwald E. Determination of fraction of left ventricular volume ejected per beat and of ventricular end-diastolic and residual volumes. Experimental and clinical observations with a precordial dilution technic. Circulation. 1962;25:674–685.

14. CIBIS-II. The Cardiac Insufficiency Bisoprolol Study II (CIBIS-II): a randomised trial. Lancet. 1999;353(9146):9–13.

15. Eichhorn EJ, Bristow MR. The Carvedilol Prospective Randomized Cumulative Survival (COPERNICUS) trial. Curr Control Trials Cardiovasc Med. 2001;2(1):20–23.

16. Straw S, McGinlay M, Witte KK. Four pillars of heart failure: contemporary pharmacological therapy for heart failure with reduced ejection fraction. Open Heart. 2021;8(1).

17. Anker SD, Butler J, Filippatos G, et al. Empagliflozin in Heart Failure with a Preserved Ejection Fraction. N Engl J Med. 2021;385(16):1451–1461.

18. Solomon SD, McMurray JJV, Claggett B, et al. Dapagliflozin in Heart Failure with Mildly Reduced or Preserved Ejection Fraction. N Engl J Med. 2022.

19. Solomon SD, Claggett B, Lewis EF, et al. Influence of ejection fraction on outcomes and efficacy of spironolactone in patients with heart failure with preserved ejection fraction. Eur Heart J. 2016;37(5):455–462.

20. Solomon SD, Vaduganathan M B LC, et al. Sacubitril/Valsartan Across the Spectrum of Ejection Fraction in Heart Failure. Circulation. 2020;141(5):352–361.

21. Pitt B, Pfeffer MA, Assmann SF, et al. Spironolactone for heart failure with preserved ejection fraction. N Engl J Med. 2014;370(15):1383–1392.

22. Solomon SD, McMurray JJV, Anand IS, et al. Angiotensin-Neprilysin Inhibition in Heart Failure with Preserved Ejection Fraction. N Engl J Med. 2019;381(17):1609–1620.

23. Cole GD, Dhutia NM, Shun-Shin MJ, et al. Defining the real-world reproducibility of visual grading of left ventricular function and visual estimation of left ventricular ejection fraction: impact of image quality, experience and accreditation. Int J Cardiovasc Imaging. 2015;31(7):1303–1314.

24. Maurer MS, Packer M. How Should Physicians Assess Myocardial Contraction?: Redefining Heart Failure With a Preserved Ejection Fraction. JACC Cardiovasc Imaging. 2020;13(3):873–878.

25. Stokke TM, Hasselberg NE, Smedsrud MK, et al. Geometry as a Confounder When Assessing Ventricular Systolic Function: Comparison Between Ejection Fraction and Strain. J Am Coll Cardiol. 2017;70(8):942–954.

26. Metra M, Pagnesi M, Claggett BL, et al. Effects of omecamtiv mecarbil in heart failure with reduced ejection fraction according to blood pressure: the GALACTIC-HF trial. Eur Heart J. 2022.

27. Sagawa K, Suga H, Shoukas AA, Bakalar KM. End-systolic pressure/volume ratio: a new index of ventricular contractility. Am J Cardiol. 1977;40(5):748–753.

28. Bozkurt B, Coats AJS, Tsutsui H, et al. Universal definition and classification of heart failure: a report of the Heart Failure Society of America, Heart Failure Association of the European Society of Cardiology, Japanese Heart Failure Society and Writing Committee of the Universal Definition of Heart Failure: Endorsed by the Canadian Heart Failure Society, Heart Failure Association of India, Cardiac Society of Australia and New Zealand, and Chinese Heart Failure Association. Eur J Heart Fail. 2021;23(3):352–380.

29. Witte KK, Patel PA, Walker AMN, et al. Socioeconomic deprivation and mode-specific outcomes in patients with chronic heart failure. Heart. 2018;104(12):993–998.

