## Supplementary material for "Cardiac contractility index identifies systolic dysfunction in preserved ejection fraction heart failure": (Table 1)

|  | All patients  (n=728) | Divided by LVEF | | | | Divided by cardiac contractility index | | | |
| --- | --- | --- | --- | --- | --- | --- | --- | --- | --- |
|  |  | Tertile 1  <46.2%  (n=242) | Tertile 2  46.2-55.1%  (n=243) | Tertile 3  >55.1%  (n=243) | *p-*value | Tertile 1  <3.65mmHg/ml/m^2^  (n=242) | Tertile 2  3.65-5.34mmHg/ml/m^2^  (n=243) | Tertile 3  >5.34mmHg/ml/m^2^  (n=243) | *p*-value |
| Demographics |  |  |  |  |  |  |  |  |  |
| Age (years) | 82.6 ± 9.2 | 81.4 ± 10.0 | 83.9 ± 8.5 | 82.6 ± 8.9 | 0.012 | 81.7 ± 10.4 | 82.9 ± 8.8 | 83.2 ± 8.2 | 0.16 |
| Male sex [n(%)] | 330 (45.3) | 148 (44.8) | 99 (30.0) | 83 (25.2) | <0.001 | 154 (46.7) | 102 (30.9) | 74 (22.4) | <0.001 |
| NYHA Class III/IV [n(%)] | 328 (45.1) | 98 (40.3) | 109 (44.9) | 121 (50.0) | 0.10 | 104 (42.8) | 103 (42.4) | 121 (50.0) | 0.17 |
| Co-morbidities |  |  |  |  |  |  |  |  |  |
| IHD [n(%)] | 210 (28.8) | 92 (37.9) | 64 (26.3) | 54 (22.3) | <0.001 | 86 (35.4) | 70 (28.8) | 54 (22.3) | 0.006 |
| Stroke/TIA [n(%)] | 62 (8.5) | 20 (8.2) | 23 (9.5) | 19 (7.9) | 0.80 | 22 (9.1) | 26 (10.7) | 14 (5.8) | 0.14 |
| Hypertension [n(%)] | 490 (67.3) | 133 (54.7) | 176 (72.4) | 181 (74.8) | <0.001 | 140 (57.6) | 166 (68.3) | 184 (76.0) | <0.001 |
| Diabetes mellitus [n(%)] | 208 (28.6) | 85 (35.0) | 64 (26.3) | 59 (24.4) | 0.023 | 85 (35.0) | 65 (26.7) | 58 (24.0) | 0.020 |
| Atrial fibrillation [n(%)] | 263 (36.1) | 87 (35.8) | 98 (40.3) | 78 (32.3) | 0.18 | 90 (37.0) | 92 (37.9) | 81 (33.5) | 0.57 |
| CKD [n(%)] | 152 (20.9) | 45 (18.5) | 58 (23.9) | 49 (20.2) | 0.33 | 53 (21.8) | 43 (17.7) | 56 (23.1) | 0.31 |
| COPD [n(%)] | 110 (15.1) | 38 (15.6) | 38 (15.6) | 34 (14.0) | 0.85 | 39 (16.0) | 42 (17.3) | 29 (12.0) | 0.23 |
| Observations |  |  |  |  |  |  |  |  |  |
| SBP (mmHg) | 140.3 ± 22.9 | 134.7 ± 23.8 | 141.8 ± 20.9 | 144.4 ± 22.9 | <0.001 | 129.5 ± 21.7 | 140.1 ± 20.2 | 151.4 ± 21.5 | - |
| Heart rate (beats/min) | 76.0 ± 16.9 | 79.1 ± 19.4 | 74.7 ± 16.4 | 74.0 ± 14.2 | 0.001 | 78.1 ± 19.3 | 74.6 ± 16.4 | 75.2 ± 14.6 | 0.049 |
| Echocardiogram |  |  |  |  |  |  |  |  |  |
| LVEDVi (ml/m^2^) | 65.0 (53.9-81.8) | 76.4 (60.1-94.0) | 62.3 (50.9-75.1) | 61.1 (52.1-72.5) | - | 86.5 (73.4-100.6) | 65.4 (56.5-74.8) | 52.1 (45.3-59.6) | <0.001 |
| LVESVi (ml/m^2^) | 31.4 (24.7-43.4) | 48.7 (36.4-65.7) | 30.2 (24.9-36.7) | 24.9 (21.2-29.6) | - | 51.2 (42.5-65.7) | 31.4 (27.5-35.2) | 23.0 (20.2-26.0) | - |
| LVEF (%) | 48.2 ± 11.6 | 34.4 ± 9.1 | 51.3 ± 2.2 | 58.9 ± 2.5 | - | 37.2 ± 11.8 | 51.6 ± 7.1 | 55.8 ± 5.0 | <0.001 |
| CCI (mmHg/ml/m^2^) | 4.55 ± 1.92 | 2.94 ± 1.30 | 4.81 ± 1.41 | 5.92 ± 1.68 | <0.001 | 37.2 ± 11.8 | 51.6 ± 7.1 | 55.83 ± 5.0 | - |
| Blood tests |  |  |  |  |  |  |  |  |  |
| Haemoglobin (g/L) | 130 (119-142) | 132 (122-144) | 129 (117-141) | 129 (119-140) | 0.045 | 128 (116-141) | 130 (119-141) | 131 (122-143) | 0.21 |
| Creatinine (mmol/L) | 82 (69-104) | 86 (74-112) | 83 (69-105) | 78 (65-94) | <0.001 | 89 (74-122.5) | 80 (68.8-100) | 77 (65-96) | <0.001 |
| Sodium | 140 (138-143) | 140 (138-142) | 141 (138-142) | 141 (138-143) | 0.063 | 140 (138-142) | 140.5 (138-143) | 141 (139-143) | 0.052 |
| Albumin (g/L) | 42 (40-44) | 42 (40-44) | 41 (39-44) | 42 (40-44) | 0.030 | 42 (39-44) | 42 (40-44) | 42 (40-44) | 0.029 |
| NT-proBNP (pg/mL) | 1066 (503.5-2570) | 2119 (810-4827) | 1003 (503-2113) | 655 (346-1344) | <0.001 | 2310 (933-5155) | 921 (486-1759) | 715 (348-1362) | <0.001 |
| HbA1c (mmol/mol) | 46 (41-56) | 47 (42-56) | 46 (40.5-56) | 45 (40-54) | 0.080 | 47 (41-58) | 45 (41-53) | 46 (40-54.5) | 0.31 |
| Medications |  |  |  |  |  |  |  |  |  |
| Beta-blocker [n(%)] | 406 (55.8) | 144 (59.3) | 131 (53.9) | 131 (54.1) | 0.41 | 135 (55.6) | 133 (54.7) | 138 (57.0) | 0.88 |
| Bisoprolol dose (mg) | 2.9 ± 3.5 | 2.8 ± 3.4 | 2.8 ± 3.5 | 3.0 ± 3.6 | 0.81 | 2.5 ± 3.2 | 3.0 ± 3.6 | 3.1 ± 3.6 | 0.10 |
| ACEi/ARB [n(%)] | 442 (60.7) | 158 (65.0) | 138 (56.8) | 146 (60.3) | 0.18 | 156 (64.2) | 134 (55.1) | 152 (62.8) | 0.089 |
| Ramipril dose (mg) | 3.4 ± 3.8 | 3.3 ± 3.7 | 3.1 ± 3.7 | 3.6 ± 4.1 | 0.39 | 3.4 ± 3.8 | 3.0 ± 3.6 | 3.7 ± 4.1 | 0.11 |
| Loop diuretic [n(%)] | 338 (46.4) | 137 (56.4) | 114 (46.9) | 87 (36.0) | <0.001 | 143 (58.8) | 103 (42.4) | 92 (38.0) | <0.001 |
| Furosemide dose (mg) | 0 (0-40) | 20 (0-40) | 0 (0-40) | 0 (0-20) | <0.001 | 20 (0-40) | 0 (0-40) | 0 (0-40) | <0.001 |
| MRA [n(%)] | 29 (4.0) | 20 (8.2) | 5 (2.1) | 4 (1.7) | <0.001 | 19 (7.8) | 6 (2.5) | 4 (1.7) | 0.001 |
