## Supplementary material for "Cardiac contractility index identifies systolic dysfunction in preserved ejection fraction heart failure": (Table 2)

|  | All patients  (n=728) | HFrEF | | HFpEF | |
| --- | --- | --- | --- | --- | --- |
|  |  | Low CCI  (n=232) | High CCI  (n=61) | Low CCI  (n=132) | High CCI  (n=303) |
| Demographics |  |  |  |  |  |
| Age (years) | 82.6 ± 9.2 | 81.1 ± 10.3^#^ | 83.2 ± 8.5 | 83.9 ± 8.4^#^ | 83.1 ± 8.4 |
| Male sex [n(%)] | 330 (45.3) | 151 (65.1)*^#^ | 23 (37.7)* | 64 (48.5)*^#^ | 92 (30.4)* |
| NYHA Class III/IV [n(%)] | 328 (45.1) | 95 (40.9) | 18 (29.5) | 55 (41.7) | 160 (52.8) |
| Co-morbidities |  |  |  |  |  |
| IHD [n(%)] | 210 (28.8) | 87 (37.5)^#^ | 25 (41.0)^#^ | 33 (25.0)^#^ | 65 (21.5)^#^ |
| Stroke/TIA [n(%)] | 62 (8.5) | 21 (9.1) | 5 (8.2) | 12 (9.1) | 24 (7.9) |
| Hypertension [n(%)] | 490 (67.3) | 123 (53.0)*^#^ | 42 (68.9)* | 94 (71.2)^#^ | 231 (76.2) |
| Diabetes mellitus [n(%)] | 208 (28.6) | 83 (35.8)^#^ | 19 (31.1) | 34 (25.8)^#^ | 72 (23.8) |
| Atrial fibrillation [n(%)] | 263 (36.1) | 81 (34.9) | 20 (32.8) | 57 (43.2) | 105 (34.7) |
| CKD [n(%)] | 152 (20.9) | 42 (18.1) | 11 (18.0) | 31 (23.5) | 68 (22.4) |
| COPD [n(%)] | 110 (15.1) | 38 (16.4) | 10 (16.4) | 22 (16.7) | 40 (13.2) |
| Observations |  |  |  |  |  |
| SBP (mmHg) | 140.3 ± 22.9 | 131.6 ± 22.8* | 149.0 ± 20.5* | 133.8 ± 20.0* | 148.0 ± 21.4* |
| Heart rate (beats/min) | 76.0 ± 16.9 | 78.7 ± 19.7^#^ | 79.5 ± 19.3^#^ | 74.4 ± 15.5^#^ | 73.8 ± 14.3^#^ |
| Echocardiogram |  |  |  |  |  |
| LVEDVi (ml/m^2^) | 65.0 (53.9-81.8) | 80.9 (66.0-97.6)* | 52.1 (45.4-57.9)*^#^ | 80.5 (69.9-89.1)* | 55.9 (48.1-63.8)*^#^ |
| LVESVi (ml/m^2^) | 31.4 (24.7-43.4) | 51.1 (40.7-67.1)*^#^ | 29.2 (24.6-32.6)*^#^ | 35.8 (32.2-41.2)*^#^ | 24.3 21.2-27.6)*^#^ |
| LVEF (%) | 48.2 ± 11.6 | 34.6 ± 9.6*^#^ | 45.0 ± 4.1*^#^ | 54.2 ± 3.5*^#^ | 56.6 ± 3.8*^#^ |
| CCI (mmHg/ml/m^2^) | 4.55 ± 1.92 | 2.64 ± 0.88*^#^ | 5.31 ± 0.77*^#^ | 3.69 ± 0.56*^#^ | 6.24 ± 1.37*^#^ |
| Blood tests |  |  |  |  |  |
| Haemoglobin (g/L) | 130 (119-142) | 131 (119-143.8)^#^ | 132 (123-144) | 125.5 (115-138.8)*^#^ | 130 (120-141)* |
| Creatinine (mmol/L) | 82 (69-104) | 86 (74-113) | 79 (73.5-100) | 83 (69.8-108)* | 78 (65-98)* |
| Sodium | 140 (138-143) | 140 (138-142)^#^ | 141 (138-142.5) | 141 (138-143)^#^ | 141 (138-143) |
| Albumin (g/L) | 42 (40-44) | 42 (39-43.8)* | 43 (41-44.5)* | 41 (38-44)* | 42 (40-44)* |
| NT-proBNP (pg/mL) | 1066 (503.5-2570) | 2235 (788-5052)*^#^ | 813 (450-1810)* | 1153 (503-2353)*^#^ | 761 (401-1409)* |
| HbA1c (mmol/mol) | 46 (41-56) | 47 (42-56) | 52 (44-62)^#^ | 44 (39-54) | 45 (40-52.8)^#^ |
| Medications |  |  |  |  |  |
| Beta-blocker [n(%)] | 406 (55.8) | 132 (56.9) | 35 (57.4) | 63 (47.7)* | 176 (58.1)* |
| Bisoprolol dose (mg) | 2.9 ± 3.5 | 2.6 ± 3.3 | 3.2 ± 3.7 | 2.6 ± 3.4 | 3.2 ± 3.6 |
| ACEi/ARB [n(%)] | 442 (60.7) | 153 (65.9)^#^ | 34 (55.7) | 69 (52.3)^#^ | 186 (61.4) |
| Ramipril dose (mg) | 3.4 ± 3.8 | 3.3 ± 3.6 | 3.2 ± 3.9 | 2.9 ± 3.7 | 3.6 ± 4.0 |
| Loop diuretic [n(%)] | 338 (46.4) | 134 (57.8)*^#^ | 24 (39.3)* | 58 (43.9)^#^ | 122 (40.3) |
| Furosemide dose (mg) | 0 (0-40) | 20 (0-40)*^#^ | 0 (0-40)* | 0 (0-40)^#^ | 0 (0-40) |
| MRA [n(%)] | 29 (4.0) | 19 (8.2) | 2 (3.3) | 3 (2.3) | 5 (1.7) |

***represents *p*<0.05 between cardiac contractility index categories within HFrEF and HFpEF groups.

^#^represents *p*<0.05 between HFrEF and HFpEF within cardiac contractility index groups.
