## Supplementary material for "Cardiac contractility index identifies systolic dysfunction in preserved ejection fraction heart failure": (Table 3)

|  |  | **Cardiac contractility index** | **LVEF** |
| --- | --- | --- | --- |
|  | **Unadjusted IRR**  **(95% CI)** | **Adjusted IRR**  **(95% CI)** | **Adjusted IRR**  **(95% CI)** |
| Age (per year) | 1.06 (1.05-1.07) | 1.06 (1.04-1.07) | 1.06 (1.04-1.07) |
| Male | 1.19 (0.99-1.42) | 1.08 (0.87-1.35) | 1.15 (0.93-1.43) |
| Ischaemic heart disease | 1.05 (0.87-1.28) | 0.94 (0.76-1.17) | 0.96 (0.77-1.19) |
| Diabetes mellitus | 1.09 (0.89-1.32) | 1.21 (0.98-1.50) | 1.22 (1.00-1.51) |
| Hypertension | 1.00 (0.83-1.21) | 1.07 (0.87-1.32) | 1.05 (0.85-1.30) |
| SBP (per mmHg) | 1.00 (0.99-1.00) | 1.00 (1.00-1.00) | 1.00 (0.99-1.00) |
| HR (per beat/min) | 1.01 (1.00-1.01) | 1.01 (1.00-1.01) | 1.01 (1.00-1.01) |
| log10 haemoglobin (per g/L) | 0.32 (0.17-0.61) | 0.87 (0.35-2.19) | 0.62 (0.25-1.51) |
| Log10 creatinine (per μmol/L) | 6.61 (3.74-11.67) | 1.59 (0.78-3.23) | 1.58 (0.77-3.24) |
| Log10 albumin | 0.00 (0.00-0.01) | 0.01 (0.00-0.16) | 0.01 (0.00-0.14) |
| Log10 NTpro-BNP | 2.19 (1.85-2.59) | 1.29 (1.02-1.62) | 1.37 (1.09-1.72) |
| Cardiac contractility index (mmHg/ml/m^2^) | | | |
| 2 | 1.56 (1.26-1.93) | 1.34 (1.03-1.75) | - |
| 4 | 1.07 (1.01-1.13) | 1.06 (1.00-1.13) | - |
| 4.43 | 1.00 | 1.00 | - |
| 6 | 0.78 (0.67-0.92) | 0.78 (0.66-0.93) | - |
| 8 | 0.54 (0.39-0.74) | 0.61 (0.43-0.85) | - |
| LVEF (%) | | | |
| 20 | 1.69 (1.24-2.29) | - | 1.27 (0.89-1.80) |
| 30 | 1.19 (0.98-1.45) | - | 1.01 (0.80-1.28) |
| 40 | 0.95 (0.79-1.13) | - | 0.89 (0.73-1.09) |
| 50 | 1.00 | - | 1.00 |
| 60 | 0.66 (0.52-0.84) | - | 0.69 (0.54-0.88) |
