## Supplementary material for "Cardiac contractility index identifies systolic dysfunction in preserved ejection fraction heart failure": (Supplementary Table 1)

| **Diagnosis** | **Frequency (n=182)** |
| --- | --- |
| Angina/ischaemic heart disease | 10 (5%) |
| Atrial fibrillation | 10 (5%) |
| Chest infection | 3 (2%) |
| Chronic obstructive pulmonary disease | 23 (13%) |
| Deconditioning | 5 (3%) |
| Dependent oedema | 6 (3%) |
| Diabetes mellitus | 2 (1%) |
| Hypertrophic obstructive cardiomyopathy | 2 (1%) |
| Hypertension | 26 (14%) |
| Lung cancer | 2 (1%) |
| Obesity | 10 (5%) |
| Right heart failure/pulmonary hypertension | 42 (23%) |
| Unascertained | 30 (16%) |
| Valvular heart disease | 11 (6%) |
